# Virtual reality intervention alleviates dyspnea in patients recovering from COVID pneumonia

**DOI:** 10.1101/2021.10.26.21265510

**Authors:** Betka Sophie, Kannape Oliver Alan, Fasola Jemina, Lance Florian, Cardin Sylvain, Schmid Aline, Similowski Thomas, Soccal Paola Marina, Herbelin Bruno, Adler Dan, Blanke Olaf

## Abstract

**Background:** Immersive virtual reality (iVR)-based digital therapeutics (DTx) are gaining clinical attention in the field of pain management. Based on known analogies between pain and dyspnea, we investigated the effects of visual-respiratory feedback, on persistent dyspnea in patients recovering from COVID-19 pneumonia.

**Methods:** We performed a controlled, randomized, single-blind, cross-over clinical study to evaluate an iVR-based intervention to alleviate dyspnea in patients recovering from COVID-19 pneumonia. Included patients reported persistent dyspnea (≥5 on a 10-point scale) and preserved cognitive function (MoCA>24). Assignment was random and concealed. Patients received synchronous (intervention) or asynchronous (control) feedback of their breathing, embodied via a gender-matched virtual body. Outcomes were assessed using questionnaires and breathing recordings. COVVR is registered with ClinicalTrials.gov (NCT04844567).

**Findings:** Study enrollment was open between November 2020 and April 2021. Twenty-six patients were enrolled (27% women; age: median=55, interquartile range (IQR)=18). Data were available for 24 of 26 patients. The median (IQR) rating on a 7-point Likert-scale of breathing comfort improved from 1(2) at baseline, to 2(1) for synchronous feedback, but remained unchanged at 1(1.5) for asynchronous feedback (p<0.05) between iVR conditions). Moreover, 91.2% of all patients were satisfied with the intervention (p<0.0001) and 66.7% perceived it as beneficial for their breathing (p<0.05). No adverse events were reported.

**Interpretation:** Based on these findings, our iVR-based DTx presents a feasible and safe respiratory rehabilitation tool that improves breathing comfort in patients recovering from COVID-19 infection presenting with persistent dyspnea. Future research should investigate the DTx’s generalizability to persistent dyspnea with other etiologies and its potential for preventing chronification.

**Funding:** Marie Sklodowska-Curie Individual Fellowship (H2020-MSCA-IF-2019 894111/ RESPVR), Bertarelli Foundation

## Introduction

Dyspnea is defined as “*a subjective experience of breathing discomfort made of various sensations that can vary in intensity*”^1^. In simpler words, dyspnea relates to the upsetting or distressing awareness of breathing activity. Beyond the symptom of cardiorespiratory dysfunction, dyspnea is a frightening and disabling experience. This is particularly true when it resists optimized treatment of the underlying condition, a situation termed “chronic breathlessness”^2^ or, more broadly, “persistent dyspnea”^3^. Persistent dyspnea deeply affects the lives of those afflicted. It profoundly deteriorates quality of life by impacting cognitive function, locomotion, and mental health^4^. Implicit to the definition of persistent dyspnea is the under-recognition of respiratory suffering as a major clinical burden. This invisibility impairs access to care and hinders the development of evidence-based targeted interventions^5^, with the current therapeutic arsenal remaining limited (e.g., opioids, or trigeminal stimulation with portable fans)^6^.

Neuroscience evidence suggests that dyspnea occurs in conjunction with the recruitment of a neural network involving the insula, dorsal anterior cingulate cortex, amygdala, and medial thalamus, sharing important pathways with other brain functions such as pain processing^10^ and bodily self-consciousness^12^. This suggests that it is relevant to target the brain to relieve dyspnea when all cardiorespiratory approaches have been exhausted ^13^. In this regard, prior interventions using immersive virtual reality (iVR)-based Digital Therapeutics (DTx), also referred to as digiceuticals^14^, have demonstrated alleviation of chronic pain in patients with complex regional pain syndrome or spinal cord injury^12,13^. In the respiratory domain, visuo-respiratory stimulation has been associated with an increased feeling of breathing control (breathing agency)^17^, a reduced negative emotional state related to experimental dyspnea^18^, as well as changes in physiological measures of breathing^11,19^.

Persistent symptoms can occur beyond the initial period of COVID-19 infection recovery and affect patients who were managed in the community or in the acute care setting^17^. Like general weakness, malaise, fatigue, and impaired concentration, dyspnea has consistently been reported in so-called long COVID cohorts with a high prevalence of around 25% (CI95 18% to 34%)^18^. In the case of persistent dyspnea, an extensive workup to identify respiratory sequelae or muscle deconditioning should be the foremost clinical preoccupation, mostly to guide the indications of pulmonary rehabilitation^19^. Yet, dyspnea can be dissociated from physiological markers such as pulmonary function tests or lung imaging, in post-COVID situations^18^ as in a more general manner ^22^. This makes treatment and even diagnosis challenging. The importance of brain mechanisms in the pathogenesis of dyspnea justifies neuroscientific approaches for its management and implies that a cognitive intervention using a neuro-rehabilitation approach could be tested to understand and alleviate this debilitating symptom.

The present COVID-19 Virtual Reality (COVVR) clinical study was performed to test the hypothesis that an iVR-based DTx would alleviate dyspnea by improving breathing comfort in patients recovering from COVID-19 pneumonia presenting with persistent dyspnea. We further evaluated participants’ perceived awareness of and agency over their breathing movements^17^. Finally, we tracked patients’ perceived benefits related to the iVR intervention and the feasibility of using COVVR in the clinic or at home.

## Method

### Study Design

A prospective controlled, randomized, single-blind, cross-over clinical study was conducted to evaluate both the efficacy and feasibility of a iVR biofeedback intervention to alleviate persistent dyspnea in patients recovering from COVID-19 pneumonia. This single-site study was carried out at the University Hospital (HUG) in Geneva, Switzerland and was approved by the Commission Cantonale d’Ethique de la Recherche de la République et Canton de Genève (2019-02360).

### Patients

Thirty-nine patients were screened by a respiratory physician (AS). Patients that scored below 25 on the Montreal Cognitive Assessment (MoCA) were excluded (N=5); N=8 refused to participate. In total, N=26 patients were enrolled. Clinical inclusion criteria were that patients) were recovering from COVID-19 pneumonia confirmed by RT-PCR for SARS-CoV-2, and ii) presented with persistent dyspnea with a self-rated intensity of five or higher (out of ten) on a visual analog dyspnea scale. The respiratory physician asked the dyspnea question as follows: “Do you have difficulty breathing?” then “On a scale of 0 to 10, with 0 being no difficulty to breathe and 10 being the worst difficulty to breathe that you can imagine, where do you rank?”. This dyspnea rating was only used as an inclusion criterion and not as an outcome. Patients had to be able to give consent and to understand and speak French or English. Patients who presented with unstable respiratory, neurological, or cardiac conditions, or psychiatric illness were excluded.

### Randomization and masking

Patients were allocated to one of two starting conditions using a randomization script (i.e., randomizing the experimental sequence for each patient, using Matlab version 2020a), before data collection. Randomization was not restricted; no stratification or minimization factors were applied. Allocation was concealed to the clinicians screening patients.

Participant masking (blinding) was achieved by keeping both the procedure and the virtual environment identical for both tested conditions. Participants were naïve to the difference in the two conditions which consisted only of a change in feedback synchrony between respiratory movements and virtual body luminance. Experimenters were not blinded, however, the instructions were only given to the patient once and applied to both conditions.

### Procedure

#### Screening

Patients were screened by a respiratory physician during morning rounds on weekdays only, except for some leave days, in the division of lung diseases of Geneva University Hospital. Once referred, it was verified that the patients met the inclusion criteria by performing an anamnestic interview as well as the MoCA. The delay between the initial screening by the physician and the inclusion by the researcher varied between 1 hour to 2 days.

#### Setup

Eligible patients were installed in a semi-seated position in their hospital bed and wore a belt-mounted linear force sensor (Go Direct® Respiration belt, Vernier, Beaverton (OR), USA) fitted on the abdomen to allow proper recording of respiratory movements. They were also equipped with a head-mounted display (Zeiss VR ONEPLUS, Oberkochen, Germany) holding a smartphone (Samsung Galaxy S8, Seoul, South Korea). The smartphone ran the VR application and connected via Bluetooth® to the respiration belt. MindMaze SA provided the hardware for the study, co-developed the application with the Laboratory of Cognitive Neuroscience at EPFL (SC, LF, HB), and deployed this on the smartphone. The application collects and processes respiratory data to render a computer-generated virtual environment in real-time.

#### Intervention Conditions

Patients were asked to look around in the VR environment and orient their gaze to a gender-matching virtual body lying on a bed next to them in a similar position as theirs (Figure 2.B) (cf. 17–19). The virtual body flashed in a waxing and waning visual effect which could be synchronous or asynchronous to the patient’s respiratory movements. In the synchronous condition, the radiance of the visual flash was maximal at the end of inspiration and minimal at the end of the expiration. In the asynchronous condition, at the end of each visual flash, a duration between 2.5 and 33.3 seconds is randomly generated for the next visual stimulation, such that the feedback is both phase-shifted and frequency-modulated with respect to the actual respiration.

**Figure 1:**
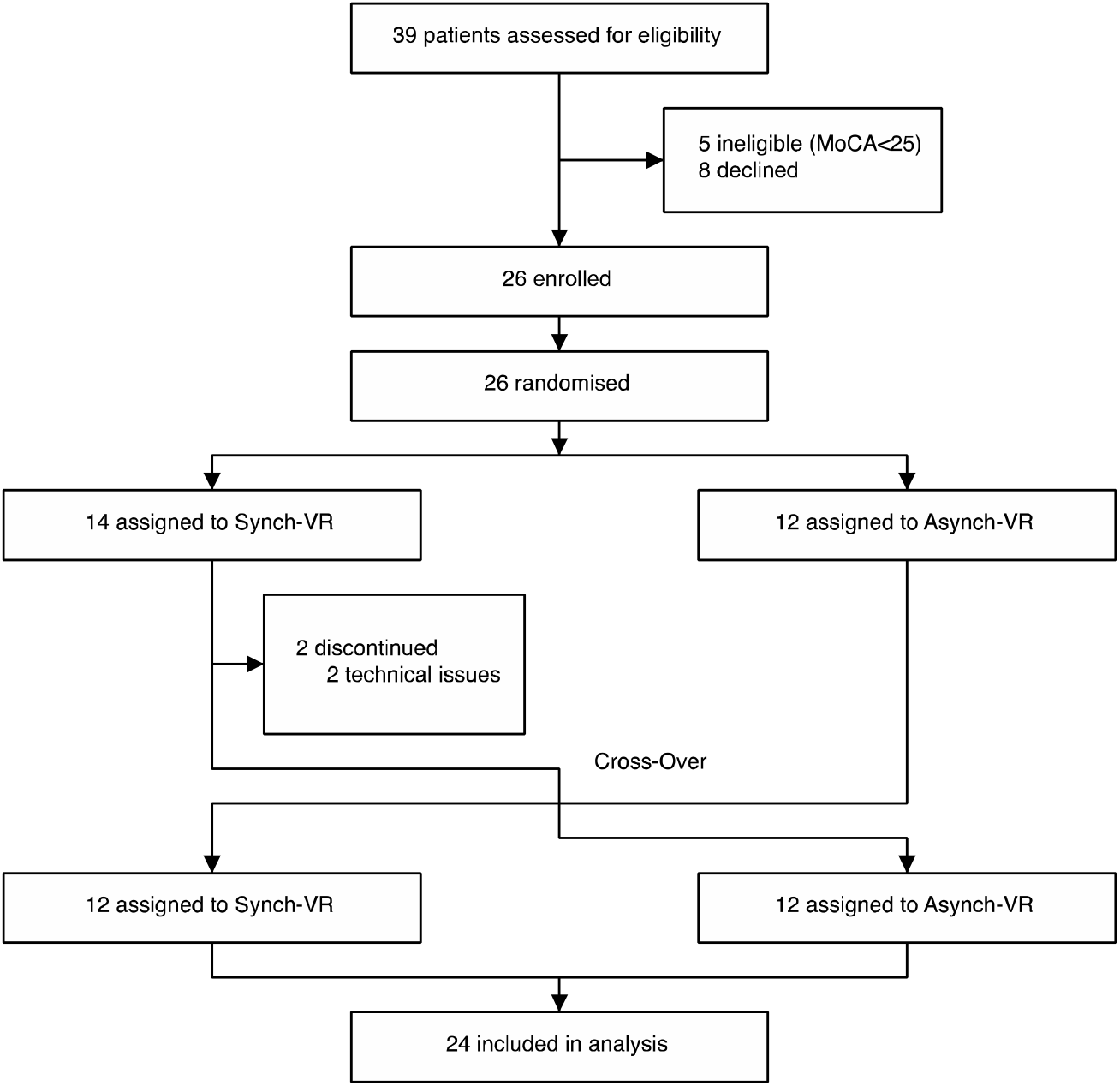
Study flow chart

**Figure 2:**
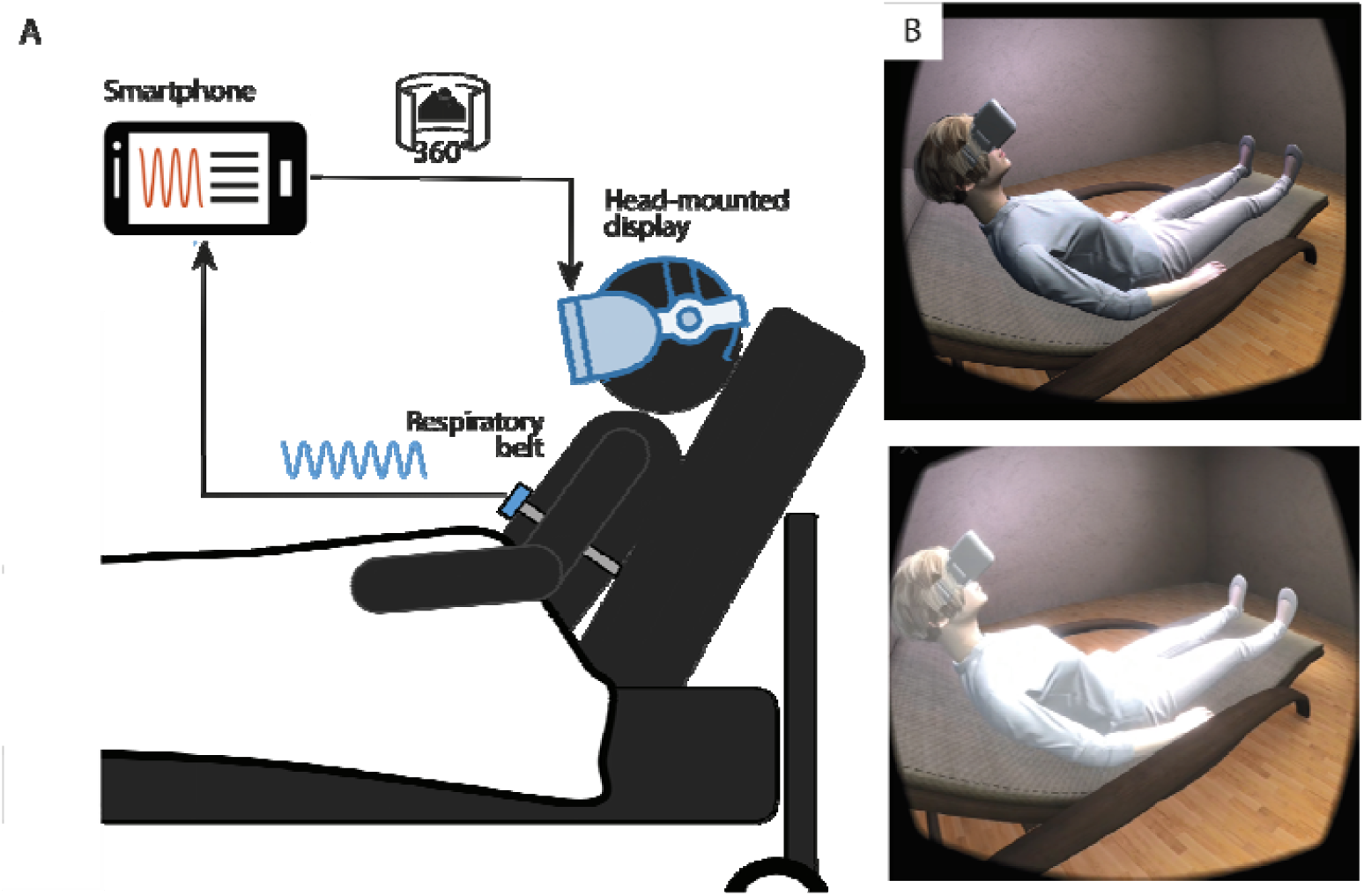
Portable setup and virtual reality feedback. (A) A respiratory belt captures the respiratory movements of the chest and sends the signal to a smartphone via Bluetooth. A custom software generates the virtual environment. (B) A matched-gender virtual body is displayed and observed by the patients by slightly turning their head to the side. The virtual body is illuminated synchronously or asynchronously with respect to the patient’s chest movements. The top image represents the end of the expiration with a low flashing intensity, while the bottom image shows the end of the inspiration corresponding to the maximal luminosity in the synchronous condition. A video of the experiment can be found in the supplementary material.

#### Intervention Procedure

Once the patient was installed, they were asked to close their eyes (no visual feedback, NVF) while their respiratory movements were recorded for two minutes. Participants were then asked to describe their current respiratory experience by answering two questions according to a 7-point Likert-scale (from -3= Strongly disagree to +3= Strongly agree): 1) *I have difficulty breathing* (breathing discomfort) and 2) *My breathing is enjoyable* (breathing comfort). This NVF condition served as the baseline assessment for the breathing (dis)comfort items and the breathing rate. For comparison the two items were also included in the post-exposure questionnaires.

After baseline assessment, participants were randomly assigned in a 1:1 ratio to receive either the sequence “synchronous/asynchronous” or “asynchronous/synchronous”. In each block, patients were first asked to look around in the virtual room, and then to orient their gaze towards the virtual body while relaxing for 5 minutes. They were not informed that the flashing of the virtual body was related to their breathing. Each block was followed by a subjective questionnaire of six items (7-point Likert scale) derived from previous visuo-respiratory studies^14–16^, (see Table 1). Question 1 evaluated the awareness of the visuo-respiratory experimental manipulation (breathing awareness)^14,16^, while question 3 pertained to the breathing agency^14–16^. Question 2 and 5 were included as control items. The breathing comfort items matched the baseline assessment (Q4 and Q6 respectively, Table 1). Finally, patients completed an *ad hoc* questionnaire to assess the acceptance and feasibility of the iVR intervention (7-point Likert scale, see supplementary section Table S1). During the entire intervention, oxygen therapy was administered through nasal cannulas to obtain a SpO2 level of 90-92%.

**Table 1.** Subjective questionnaire items

| Items |  | Domain |
| --- | --- | --- |
| Q1 | It seemed as if the flashing was my respiration | Breathing awareness |
| Q2 | It seemed as if I had three bodies | Control |
| Q3 | I felt as if the virtual body was breathing with me | Breathing agency |
| Q4 | I had difficulty breathing | Discomfort |
| Q5 | I felt as if the virtual body was drifting with the flashing | Control |
| Q6 | My breathing was enjoyable | Comfort |

### Outcomes

Efficacy and feasibility were defined as primary outcomes of the iVR intervention. Efficacy was evaluated based on subjective feedback by the patients regarding their breathing comfort and discomfort (Q4 and Q6 of the above Table 1). Feasibility was evaluated using a feedback questionnaire, alongside open feedback. Agreement with the questionnaire items indicates better feasibility, acceptance, and perceived outcome.

Secondary outcome measures included respiratory parameters as well as the subjective reports of breathing awareness and agency. Both respiratory rate (RR, breaths per minute) and respiratory rate variability (RRV, using inter-breath intervals) were measured using the respiration belt. RR and RRV were compared across the baseline and two intervention conditions. Breathing awareness and agency were evaluated using a 7-point Likert scale where agreement indicates stronger embodiment of the feedback.

### Statistical Analysis

Based on previous work on breathing agency^18^, a sample size of 21 patients was estimated, using a two-sided paired t-test with an effect size of 0.65, alpha of 0.05 and a power of 0.8 to demonstrate a breathing agency difference of 0.5 point measured on a 7-point Likert-scale from -3 to 3-in subjective ratings between the two experimental conditions (i.e., synchronous and asynchronous). The effect of synchrony on each measure was assessed using a linear mixed-effects model with a random intercept for each patient. In addition to the experimental condition (synchronous vs asynchronous condition), each model also included the experimental sequence (starting the experiment with synchronous or asynchronous condition) and the interaction between the experimental sequence and the experimental condition as fixed effects. The statistical significance of the interactions was assessed using the likelihood ratio test. All p-values were two-sided and statistical significance was set at a p-value of 0.05.

Median, interquartile range (IQR), and rating frequency (in%) were computed for each feasibility item. To ensure clarity, observed percentages for ratings from 1 = Agree to 3 = Strongly agree were grouped, indicating overall agreement with the statement. A one-sided, one-sample t-test was used to determine if the mean of ratings was significantly greater than zero, indicating that, at least, the majority of patients were agreeing with the statement.

As head-mounted displays are widely used in clinical and healthy populations no specific safety analysis was performed within the scope of this study. There is no evidence that using HMDs carry risks beyond those of CRT screens (e.g., with respect to binocular vision or photosensitive epilepsy). As patients remained seated during the intervention there was no risk of falling or collisions.

All analyses were performed using R (version 4.1.0) and Matlab (version 2020a).

## Results

Patient enrollment, randomization, and testing took place at the division of Pneumology at Geneva University Hospital between November 2020 and April 2021. Twenty-six patients were randomly assigned either to the “asynchronous/synchronous” sequence (N=12) or the “synchronous/asynchronous” sequence (N=14). At the time of database lock in May 2021, data was available for all except two (7.7 %) of 26 patients (Figure 1).

Subjective and physiological measures are reported in Table 2. At baseline, the median (and IQR) breathing comfort rating was 1(2) and the mean breathing discomfort rating was 1(3). Median values and interquartile ranges for each experimental condition, in function of the experimental sequence are provided in supplementary Table S2.

**Table 2.** Characteristics of the patients at randomisation in the intent to treat population. Data are presented as n (%) or median/IQR(Range). IQR: interquartile range. *Data were missing for some patients; the denominator in the asynchronous group was 10.

|  | **Total (n=26)** | **Synchronous first (n=14)** | **Asynchronous first (n=12)** |
| --- | --- | --- | --- |
| **Gender** |  |  |  |
| Male n (%) | 19 (73%) | 11 (79%) | 8 (67%) |
| Female n (%) | 7 (27%) | 3 (21%) | 4 (33%) |
| **Age** | 55/18 (35-81) | 55/18 (38-81) | 56.5/16.75 (35-73) |
| **MoCA** | 27/3 (25-30) | 27.5/1.75 (25-30) | 27/4.24 (25-30) |
| **SpO2 on oxygen therapy\*** | 94/4.3 (90-98) | 95/5.5 (90-98) | 92.5/4 (91-96) |
| **Oxygen flow (l/m)\*** | 1/3 (0-8) | 2/2 (0-8) | 0.25/1.75 (0-4) |
| **Heart rate (bpm)\*** | 74.5/22 (52-108) | 79.5/24.5 (62-108) | 70.5/11.5 (62-92) |
| **Days since first symptom onset \*** | 17/22 (3-39) | 14/21 (3-39) | 18.5/16.25 (6-37) |
| **Contagious at time of testing n (%)** | 16 (62%) | 10 (71%) | 7 (58%) |
| **Breathing comfort** | 1/2 (-3-2) | 0/2 (-3-2) | 1/3.25 (-2-2) |
| **Breathing discomfort** | 1/3 (-3-2) | 1/3 (-3-2) | 0/3.25 (-3-2) |
| **Respiratory rate (bpm)** | 20.7/10.0 (6.8-34.6) | 20.7/10.0 (15.2-31.5) | 22.3/10.0 (15-34.6) |
| **Respiratory rate variability (bpm)** | 3.2/1.9 (0.8-11.5) | 3.2/2.0 (1.2-11.5) | 3.1/1.7 (0.8-4.7) |

**Table 3.** Results summary for the subjective and physiological measures. #: depicts the mean difference between synchronous and asynchronous conditions, regardless of the sequence, as well as its CI estimated by the linear mixed model (the p-value corresponds to the test of this difference being equal to zero); ¶: depicts the mean difference between synchronous and asynchronous conditions and its CI estimated by the linear mixed model for the experimental sequence “Asynchronous first” (the p-value corresponds to the result of the interaction test); +: depicts the mean difference between synchronous and asynchronous conditions and its CI estimated by the linear mixed model for the experimental sequence “Synchronous first” (the p-value corresponds to the result of the interaction test). CI: confidence interval, LB: Lower Bound, UB: Upper Bound. Subjective ratings were measured using a 7-point Likert scale with -3 = Strongly disagree, -2 = Disagree; -1 = Somewhat disagree; 0 = Neither agree nor disagree; 1 = Somewhat agree; 2 = Agree; 3= Strongly agree.

| **Breathing comfort** | **beta** | **95% CI LB** | **95% CI UP** | **P-value** |
| --- | --- | --- | --- | --- |
| Main effect of Synchrony<sup>#</sup> | 0.542 | 0.046 | 1.037 | 0.033 |
| Asynch First<sup>¶</sup> | 0.25 | -0.627 | 1.127 | 0.223 |
| Synch First<sup>+</sup> | -0.583 | -1.544 | 0.378 |  |
| **Breathing discomfort** | **beta** | **95% CI LB** | **95% CI UP** | **P-value** |
| Main effect of Synchrony<sup>#</sup> | -0.5 | -1.064 | 0.064 | 0.080 |
| Asynch First<sup>¶</sup> | -1.333 | -2.260 | -0.407 | 0.221 |
| Synch First<sup>+</sup> | -0.667 | -1.760 | 0.427 |  |
| **Agency** | **beta** | **95% CI LB** | **95% CI UP** | **P-value** |
| Main effect of Synchrony<sup>#</sup> | 1.583 | 0.335 | 2.832 | 0.014 |
| Asynch First<sup>¶</sup> | -0.667 | -1.922 | 0.589 | 0.336 |
| Synch First<sup>+</sup> | 1.167 | -1.285 | 3.618 |  |
| **Awareness** | **beta** | **95% CI LB** | **95% CI UP** | **P-value** |
| Main effect of Synchrony<sup>#</sup> | 2.167 | 1.068 | 3.266 | < 0.0001 |
| Asynch First<sup>¶</sup> | 0.167 | -0.894 | 1.227 | 0.064 |
| Synch First<sup>+</sup> | 2 | -0.121 | 4.121 |  |
| **Control (Q2)** | **beta** | **95% CI LB** | **95% CI UP** | **P-value** |
| Main effect of Synchrony<sup>#</sup> | 0.042 | -0.042 | 0.125 | 0.312 |
| Asynch First<sup>¶</sup> | -3.000 | -3.135 | -2.865 | 0.302 |
| Synch First<sup>+</sup> | 0.083 | -0.080 | 0.246 |  |
| **Control (Q5)** | **beta** | **95% CI LB** | **95% CI UP** | **P-value** |
| Main effect of Synchrony<sup>#</sup> |  |  |  | Did not converge as data are similar in both conditions |
| Asynch First<sup>¶</sup> |  |  |  |  |
| Synch First<sup>+</sup> |  |  |  |  |
| **Respiration Rate** | **beta** | **95% CI LB** | **95% CI UP** | **P-value** |
| Main effect of Synchrony<sup>#</sup> | -0.275 | -1.748 | 1.198 | 0.704 |
| Asynch First<sup>¶</sup> | 23.346 | 19.460 | 27.231 | 0.053 |
| Synch First<sup>+</sup> | 2.685 | -0.041 | 5.410 |  |
| **Respiration Rate Variability** | **beta** | **95% CI LB** | **95% CI UP** | **P-value** |
| Main effect of Synchrony<sup>#</sup> | -0.295 | -0.779 | 0.190 | 0.222 |
| Asynch First<sup>¶</sup> | 4.594 | 3.344 | 5.843 | 0.810 |
| Synch First<sup>+</sup> | -0.114 | -1.082 | 0.854 |  |

Regarding the primary outcome breathing comfort, we observed that the median (and IQR) rating improved from 1(1.5) during the asynchronous condition and 2(1) during the synchronous condition, with an estimated difference of 0.54 (95% CI 0.05-1.04, p<0.05,Figure 3.A). Moreover, post-hoc paired one-sided t-tests, confirmed a significant difference between breathing comfort ratings during the intervention (synchronous) condition compared to baseline. Such difference was absent for the control (asynchronous) condition, excluding a mere effect of VR distraction (see supplementary section for statistical details). For the assessment of discomfort, even though a similar trend was observed in the data, no significant main effect of experimental condition was observed (Figure 3.D). The experimental sequence had no significant effect on breathing comfort or discomfort ratings.

**Figure 3.**
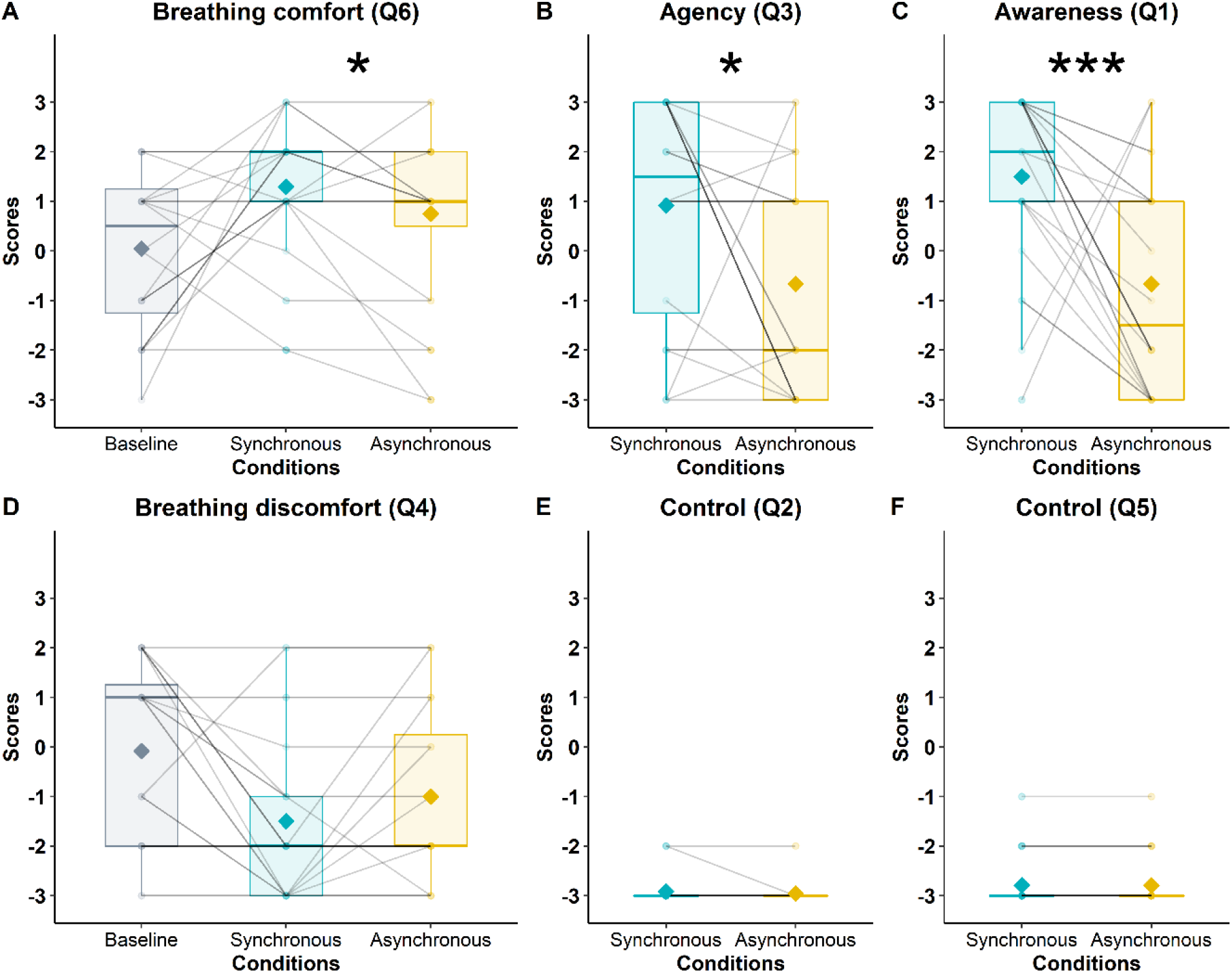
Breathing comfort (A), Agency (B), Awareness (C), Breathing discomfort (D) and control items (E&F) test results. **(A-C)** Subjective measures for which the main effect of the experimental manipulation was significant. *p<.05; ***p<.001 **(D-F)** Subjective measures for which the main effect of the experimental manipulation was not significant.

Feasibility ratings, the co-primary outcome, are depicted in Figure 4. Most of the patients (91.2%) were satisfied with the intervention (Satisfaction: median(IQR)=2(2); t=5.20, p<0.0001, 95% CI 1.17 to inf). In addition, 66.7% rated the iVR intervention as beneficial for their breathing (Respiratory benefit: 1(2.25), t=1.81, p<0.05, 95% CI 0.04 to inf). Half of the patients reported that it made them feel better (Well-being benefit: 0.5(4), t=0.36, p>0.05, 95% CI -0.64 to inf), and a further 45.8 % indicated that they would like to continue using the device during their recovery (Rehabilitation: 0(4), t=0.10, p>0.05, 95% CI -0.67 to inf) and at home (Home use: -1(4.25), t=-0.74, p>0.05, 95% CI -1.11 to inf). Finally, 37.5 % would have liked to use the intervention earlier during their stay at the hospital (Hospital use: 0(2.25), t=-0.22, p>0.05, 95% CI -0.74 to inf). Descriptive statistics and statistical tests are described in the supplementary section (Table S3 and Figure S2).

**Figure 4.**
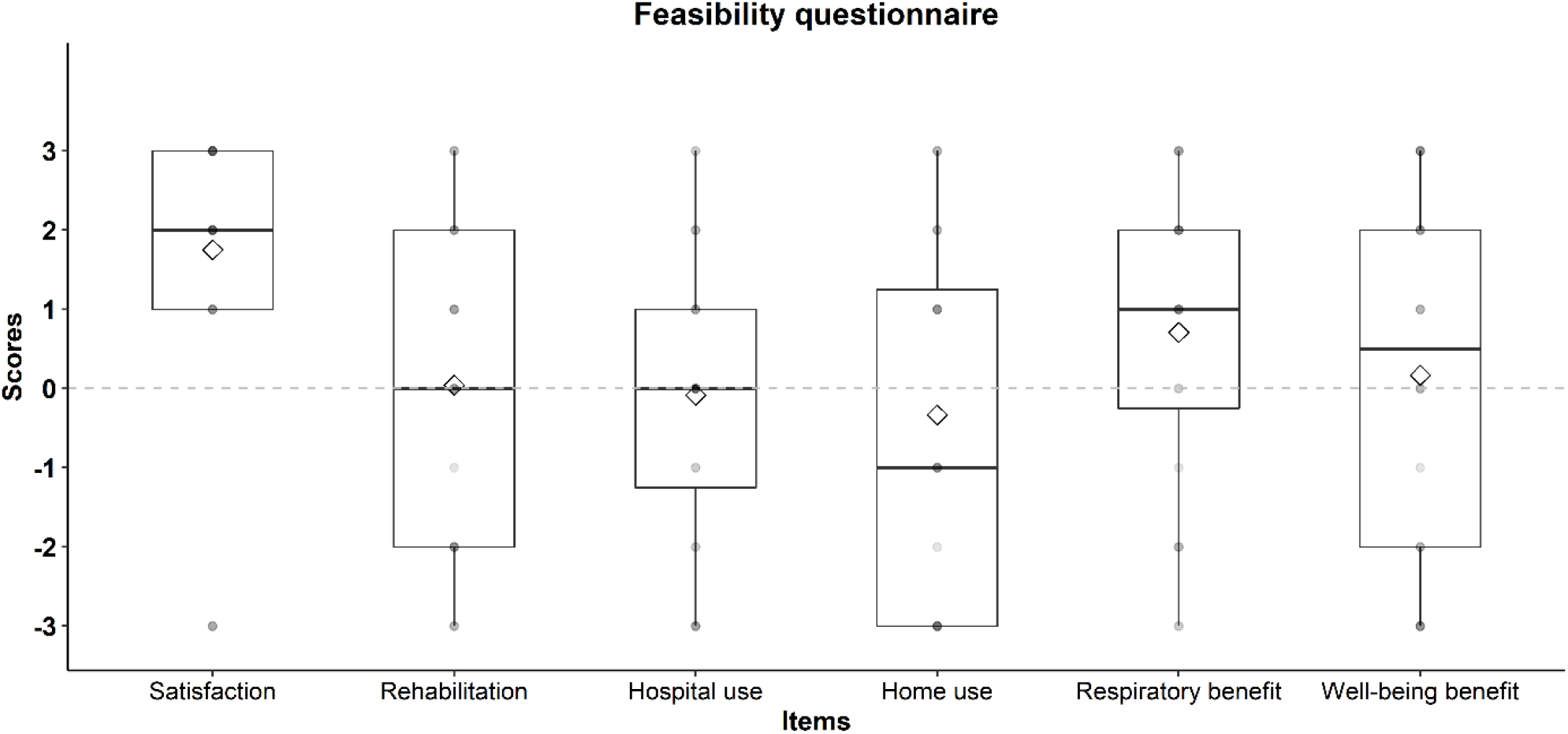
Feasibility scores for all items. The boxplots are depicting subjects’ ratings for feasibility items. The thick line within a box plot represents the median, the diamond represents the mean, the upper boundary of the box indicates the 25th percentile (Q1) and lower boundary the 75th percentile (Q3). The whiskers above and below the box indicate the minimal and maximal values (Q1 – 1.5*IQR and Q3 + 1.5*IQR respectively), while points above the upper or below the whiskers indicate outliers. Subjective ratings were measured using a 7-point Likert scale with -3 = Strongly disagree, -2 = Disagree; -1 = Somewhat disagree; 0 = Neither agree nor disagree; 1 = Somewhat agree; 2 = Agree; 3= Strongly agree.

The secondary outcome measures of this study included the subjective ratings for breathing awareness and agency and the physiological measures. The median (and IQR) breathing agency rating increased from -2(4) during the asynchronous condition to 1.5(4.25) during the synchronous condition, with an estimated difference of 1.58 (95% CI 0.34 to 2.83, p<0.05, Figure 3.B). The median breathing awareness rating increased from -1.5(4, asynchronous) to 2(2, synchronous), with an estimated difference of 2.17 (95% CI 1.07 to 3.27, p<0.0001, Figure 3.C). Neither control item differed between conditions (“*It seemed as if I had three bodies*”, Figure 3.E, “*I felt as if the virtual body was drifting with the flashing*”, Figure 3.F). Respiration rate and its variability did not differ between experimental conditions. The order of conditions did not significantly affect any of the secondary outcomes (see Figure S1).

The boxplots are depicting subjects’ ratings during asynchronous condition compared to the synchronous condition, independent of experimental sequence. The thick line within a box plot represents the median, the diamond represents the mean, the upper boundary of the box indicates the 25th percentile (Q1) and lower boundary the 75th percentile (Q3). The whiskers above and below the box indicate the minimal and maximal values (Q1 – 1.5*IQR and Q3 + 1.5*IQR respectively), while points above the upper or below the whiskers indicate outliers. Subjective ratings were measured using a 7-point Likert scale with -3 = Strongly disagree, -2 = Disagree; -1 = Somewhat disagree; 0 = Neither agree nor disagree; 1 = Somewhat agree; 2 = Agree; 3= Strongly agree.

## Panel: Research in context

### Evidence before the study

We searched MEDLINE (PubMed) to identify relevant studies investigating the relationships between respiration, persistent dyspnea/ chronic breathlessness, Virtual Reality and COVID-19 infection. We also explored studies/randomized controlled trials (RCTs) of immersive VR-based DTx aimed at alleviating pain/dyspnea. To maximize sensitivity, we also searched for citations in Google Scholar, Scopus, and Web of Science. Search terms included “COVID-19”, “coronavirus”, “breathlessness”, “dyspnea”, “virtual reality”, “breathing”, “respiration”, “rehabilitation”, “pain” and “immersive digiceuticals”, either separately or in combination. In previous work in healthy populations, the manipulation of visuo-respiratory signals using VR was related to an increased feeling of breathing control, reduced negative emotion during induced dyspnea and changes in physiological measures of breathing signal. Dyspnea is poorly associated with physiological impairment measured by pulmonary function tests or lung imaging in long COVID-19, which also implies that the brain could be a possible target for a neurorehabilitation intervention. Alleviating breathlessness in patients recovering from COVID-19 pneumonia requires a clinically proven, easily adaptable, and inexpensive intervention.

### Added value of this study

Previous iVR interventions manipulating visual cardiac feedback led to pain alleviation in clinical populations such as complex regional pain syndrome or spinal cord injury. In the respiratory domain, the manipulation of visuo-respiratory stimulations in healthy subjects has been associated with reduced negative emotional state related to dyspnea, with changes in respiratory signals (rate, tidal volume variability and respiration amplitude), and with increased breathing agency (i.e., the feeling of sensing one’s breathing command), but no data are available in patients. In our COVVR controlled, randomized, single-blind cross-over clinical study of 26 patients presenting with persistent dyspnea after COVID-19 infection, a 5-minute synchronous visuo-respiratory stimulation improved breathing comfort versus baseline and a matched control condition (also consisting of visuo-respiratory stimulation). These effects were not caused by extraneous differences in breathing frequency and variability. Most patients (22 out of 24) were satisfied by the intervention and two-thirds rated the iVR intervention as beneficial to improving this debilitating symptom.

### Implications of all the available evidence

There is a scarcity of evidence-based treatments to alleviate persistent dyspnea and prevent its chronification. The visuo-respiratory iVR-based intervention presented here improved breathing comfort in patients recovering from COVID-19 suffering from persistent dyspnea. Our iVR DTx may therefore be considered as a safe and inexpensive neurorehabilitation tool to complement existing interventions both for in-patient and out-patient populations. It may provide an additional treatment and assessment option while minimizing risk of transmission and avoiding documented side effects associated to opioid treatment. To understand the generalizability of the iVR DTx, COVVR should be evaluated in populations suffering from persistent dyspnea caused by other etiologies while longitudinal studies should be conducted to optimize dosage and evaluate its efficacy.

## Discussion

In this study, COVVR, an iVR-based immersive DTx, improved breathing comfort in patients with persistent dyspnea recovering from COVID-19 pneumonia. Persistent dyspnea, a common but underreported condition, is defined as the breathlessness reported by patients despite receiving state-of-the-art treatment of their respiratory condition, and leads to major disabilities impacting cognition, locomotion, and mental health^4,5,26^. COVVR may therefore provide an additional non-invasive and non-pharmacological tool for aiding patient recovery and satisfaction, with the potential of alleviating some of the burden on the health system given the increasing numbers of patients experiencing persistent dyspnea after COVID-19 infection.

Patients reported a significant improvement in breathing comfort after a relatively short exposure to synchronous visuo-respiratory COVVR stimulation compared to the asynchronous control condition and compared to their baseline breathing comfort. Our results extend recent observations in chronic pain studies^12^ to patients with persistent dyspnea. This previous iVR work indicated the value of personalized iVR based on cardio-visual^9,12^, somatosensory-visual^13^, and in the present study respiratory-visual feedback (see ^14–16^). Such iVR studies using multisensory bodily stimulations (including the present VR protocol), differ from previous interventions focused on using (1) immersive or non-immersive VR as a distraction tool ^21^ or (2) the more recent efforts to digitizing patient education and cognitive behavioral therapy^22^; they differ by being designed to impact the central processing of nociceptive and respiratory signals respectively. The specificity of these personalized iVR interventions, including COVVR, is highlighted by the strictly matched control condition (i.e. asynchronous condition) differing only in cardiac or respiratory synchrony, while being identical in all other aspects of VR exposure (i.e., presence of a virtual body animated by patient’s own breathing, total duration of breathing sequence, identical 3D virtual environment, etc.). This is markedly different from the more commonly applied, non-immersive VR-stimulations prevalent in medical research^21,22^.

Next to the positive primary outcome, a similar beneficial effect of the intervention was observed for breathing agency, that is, the feeling of being in control of one’s breathing. Patients reported a stronger sense of control over their breathing for synchronous feedback as well as maintained awareness of their breathing movements. Patients further reported a global satisfaction regarding the VR intervention and, more importantly, indicated that the iVR feedback improved their breathing. The COVVR study extends respiratory iVR studies in healthy individuals that have demonstrated increased breathing agency^17^, changes in tidal volume variability^19^, and translates the approach to the bedside. Monitoring these markers as well as the patient’s emotional state^18^, may be instrumental to decreasing dyspnea-related anxiety and understanding its chronification.

While synchronous visuo-respiratory stimulation improved breathing comfort, we did not observe a significant alleviation of breathing discomfort in this initial cohort. One reason is that, although participants reported persistent dyspnea with a self-rated intensity of five or higher when screened by the respiratory physician, their agreement with the discomfort item just prior to the VR intervention was quite low, cf. baseline ratings in figure 3D, potentially indicative of a white coat effect. Nonetheless, discomfort ratings followed the same pattern observed for breathing comfort, suggesting that this item should be re-evaluated in a larger sample. Another possible explanation could also lie in the semantics of the chosen items. The breathing comfort items “*My breathing was enjoyable*” taps into affective processes while the breathing discomfort items “*I had difficulty breathing*” could rather tap into sensory-cognitive processes.

Mounting evidence using functional neuroimaging suggests that patients suffering from persistent dyspnea may exhibit “hypersensitivity” to afferent respiratory signals as a result of learned expectations^23^. Perception and anticipatory processes of dyspnea are known to share breathing control mechanisms in the brainstem and the insular cortex ^23^. Consequently, once treatment of the underlying respiratory pathophysiology has been optimized, these neurorespiratory mechanisms should be considered as potential targets for pharmacological and non-pharmacological interventions for dyspnea^13^. Pharmacological treatments have been shown to be useful: low dose oral sustained-release morphine administered for persistent dyspnea is associated with improved health status in COPD without affecting PaCO2 or causing serious side-effects (especially in patients with mMRC stage 3-4)^27^. Pulmonary rehabilitation, an evidence-based multidisciplinary non-pharmacological intervention has also been shown to modify neural responses to learned breathlessness associations, likely due to central desensitization to dyspnea^25^. While not directly investigating neurorespiratory mechanisms, the present iVR paradigm, using carefully controlled visuo-respiratory conflicts, not only introduces a new complementary rehabilitation intervention but may help identify subjective (agency, awareness, dis/comfort) and physiological (breathing rate and variability) markers of “hypersensitivity”, based on perceptual and anticipatory brain processes of dyspnea.

DTx are becoming popular in the field of chronic pain management^21^. Dyspnea and pain share several similarities^7^. They engage similar brain networks^10^, are best characterized by multidimensional models^26^, and both respond to opioid treatment. As the global COVID-19 pandemic has progressed, a significant proportion of patients experience prolonged symptoms beyond the initial period of acute infection, such as persistent dyspnea^18^. The increasing number of patients isolated for prolonged periods has stressed an urgent need to develop multidisciplinary rehabilitation strategies that can be individualized and adapted to accommodate patients’ needs^27^. Given our findings, we propose that our iVR DTx is a feasible and safe neuro-rehabilitation tool that could be considered to improve breathing comfort in patients experiencing persistent dyspnea after COVID-19 infection. As our iVR DTx intervention involves neurorespiratory processes its use could further be extended to persistent dyspnea with other etiologies. Offering a DTx that can readily be adapted for home-use may be particularly relevant at a time when over 40% of adults are estimated to avoid medical care because of COVID-19-related concerns^28^.

Our study comes with certain limitations. For one, although it is based on an adequate power calculation for a proof-of-concept study, our results stem from a small sample. For another, almost half of the patients who were selected as being severely dyspneic (i.e., visual analogic dyspnea scale ≥ 5) reported low agreement with the breathing discomfort item, at baseline. This may be due to the delay between the initial screening and the start of the intervention or the fact that the former was completed by the respiratory physician and the latter by a researcher. While we here focused on a homogeneous population of patients recovering from COVID-19 infection, our intervention should be tested in a larger cohort of patients with persistent dyspnea to improve generalizability. Another important unanswered question is whether the effects observed after this short intervention can persist when patients are off-treatment. Nonetheless, our data demonstrate the value and adaptability of a personalized iVR DTx based on off-the-shelf hardware for clinical use. Longer-term dyspnea studies should, aside from the primary health and patient satisfaction outcomes, assess the economics of implementing this iVR DTx as has been done for pain therapy in hospitalized patients ^29^

In conclusion, our study shows that a short exposure to an iVR-based digital therapeutic can improve breathing comfort and breathing control in patients recovering from COVID-19 pneumonia. Global satisfaction and respiratory benefit from the patients are reported, attesting to the feasibility of the present intervention. Although more clinical data are needed, our iVR-based DTx may become a key factor of the multi-dimensional treatment of persistent dyspnea.

## Data Availability

All data produced in the present study are available upon reasonable request to the authors

## Authors’ contributions

Every author named at the start of the article contributed to the study: Conceptualization (AD, BO, BS, HB, KO, ST), Data curation (BS, FJ), Formal Analysis (FJ, BS), Funding acquisition (AD, BO, BS), Investigation (FJ, SA), Methodology (AD, BO, BS, FJ, HB, KO), Project administration (AD, FJ, SA), Resources, Software (CS, HB, LF), Supervision (AD, BO, BS, KO), Visualization (BS, FJ), Writing – original draft (BS, FJ), Writing – review & editing (AD, BO, BS, CS, HB, KO, ST). All authors confirm that they had full access to all the data in the study and accept responsibility to submit for publication. FJ and BS verified the data.

## Data sharing

The anonymised patient data will be shared, while safeguarding the privacy of patients, via the LNCO data repository (https://gitlab.epfl.ch/lnco-public).

## Acknowledgements

We thank Dr Elise Dupuis-Lozeron and Dr Fosco Bernasconi for their statistical advice.

## Ethics committee approval

This single-site study was carried out at the University Hospital (HUG) in Geneva, Switzerland and was approved by the Commission Cantonale d’Ethique de la Recherche de la République et Canton de Genève (2019-02360).

