## Supplementary material for "Virtual reality intervention alleviates dyspnea in patients recovering from COVID pneumonia"

| Items | | Domain |
| --- | --- | --- |
| Q1 | Did you enjoy the VR experience? | Satisfaction |
| Q2 | Would you like to continue using the device during your recovery? | Rehabilitation |
| Q3 | Would you have liked to use this earlier during your stay at the hospital? | Hospital Use |
| Q4 | Would you like to continue using the device at home? | Home Use |
| Q5 | Do you think the VR feedback improved your breathing? | Respiratory benefit |
| Q6 | Did the VR feedback make you feel better? | Well-being benefit |

Table S1 Feasibility questionnaire

### Supplementary Results

#### Means and standard deviations in function of the experimental sequence

|  | **Asynchronous Condition** | | | | **Synchronous Condition** | | | |
| --- | --- | --- | --- | --- | --- | --- | --- | --- |
|  | **Asynchronous first (n=12)** | | **Synchronous first (n=12)** | | **Asynchronous first (n=12)** | | **Synchronous first (n=12)** | |
| **Measures** | *Median* | *IQR* | *Median* | *IQR* | *Median* | *IQR* | *Median* | *IQR* |
| **Comfort** | 1 | 3 | 2 | 1 | 2 | 1 | 2 | 1 |
| **Discomfort** | -2 | 2 | -1 | 2 | -2 | 2 | -2 | 1 |
| **Agency** | -2 | 3,5 | -0,5 | 4 | 1,5 | 5 | 1,5 | 2 |
| **Awareness** | 1 | 3 | -2 | 3 | 3 | 3 | 2 | 2 |
| **Control (Q2)** | -3 | 0 | -3 | 0 | -3 | 0 | -3 | 0 |
| **Control (Q5)** | -3 | 0 | -3 | 0 | -3 | 0 | -3 | 0 |
| **Respiratory rate** | 20,33 | 6,34 | 21,66 | 5,22 | 20,33 | 10,87 | 21,10 | 7,69 |
| **Respiratory rate Variability** | 4,26 | 2,43 | 4,91 | 3,35 | 4,33 | 1,73 | 4,07 | 2,86 |

Table S2 Medians and interquartile ranges for the asynchronous and asynchronous conditions, in function of the experimental sequence.

#### Breathing comfort - Tests against baseline

Using post-hoc paired one-sided t-tests, we found a significant difference between breathing comfort ratings during the synchronous condition compared to baseline (Difference: 1.25±0.431, t = 2.901, p < 0.01, 95% CI 0.511 to inf). This was not observed between breathing comfort ratings during the asynchronous condition compared to baseline (Difference: 0.708±0.547, t = 1.296, p > 0.05, 95% CI -0.229 to inf), excluding a mere effect of VR distraction.

#### Additional figures

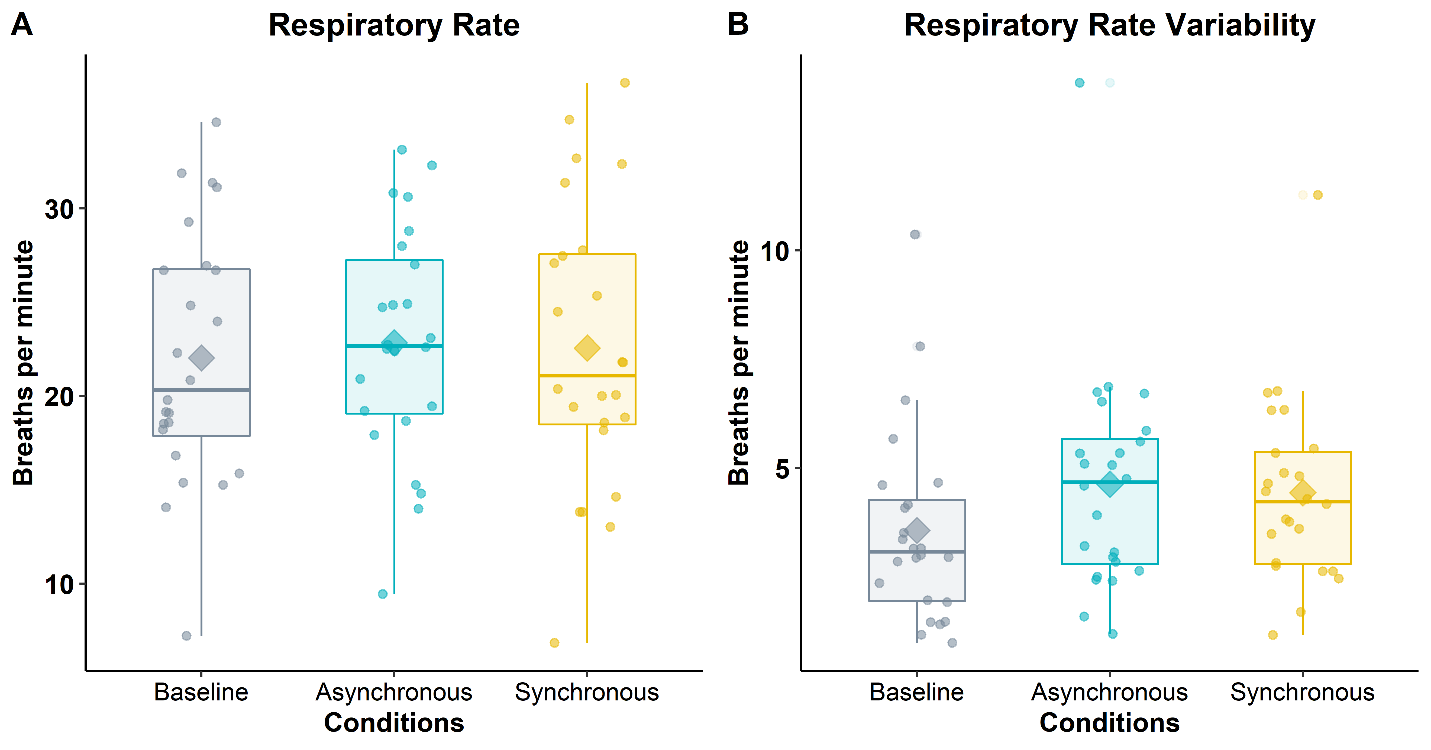

Figure S1 Objective measures for which the main effect of the experimental manipulation was not significant. The boxplots depicting subjects’ physiology signal during asynchronous condition compared to the synchronous condition, independent of experimental sequence. The thick line within a box plot represents the median, the diamond represents the mean, the upper boundary of the box indicates the 25th percentile (Q1) and lower boundary the 75th percentile (Q3). The whiskers above and below the box indicate the minimal and maximal values (Q1 – 1.5*IQR and Q3 + 1.5*IQR respectively), while points above the upper or below the whiskers indicate outliers. Subjective ratings were measured using a 7-point Likert scale with -3 = Strongly disagree, -2 = Disagree; -1 = Somewhat disagree; 0 = Neither agree nor disagree; 1 = Somewhat agree; 2 = Agree; 3= Strongly agree.

#### Feasibility

| **Items** | **Mean** | **SD** | **Median** | **IQR** | **t** | **df** | **p-value** | **CI LB** | **CI UB** |
| --- | --- | --- | --- | --- | --- | --- | --- | --- | --- |
| Satisfaction | 1.75 | 1.649 | 2.00 | 2.00 | 5.201 | 23.000 | 0.000 | 1.173 | inf |
| Rehabilitation | 0.042 | 2.032 | 0.00 | 4.00 | 0.100 | 23.000 | 0.460 | -0.669 | inf |
| Usage at the hospital | -0.083 | 1.863 | 0.00 | 2.25 | -0.219 | 23.000 | 0.586 | -0.735 | inf |
| Usage at home | -0.333 | 2.22 | -1.00 | 4.25 | -0.736 | 23.000 | 0.765 | -1.110 | inf |
| Respiratory benefit | 0.708 | 1.922 | 1.00 | 2.25 | 1.806 | 23.000 | 0.042 | 0.036 | inf |
| Well-being benefit | 0.167 | 2.297 | 0.50 | 4.00 | 0.355 | 23.000 | 0.363 | -0.637 | inf |

Table S3 Descriptive statistics and statistical tests of the feasibility items. Ratings were measured using a 7-point Likert scale with -3 = Strongly disagree, -2 = Disagree; -1 = Somewhat disagree; 0 = Neither agree nor disagree; 1 = Somewhat agree; 2 = Agree; 3= Strongly agree. SD = standard deviation, IQR = Interquartile range, df = degree of freedom

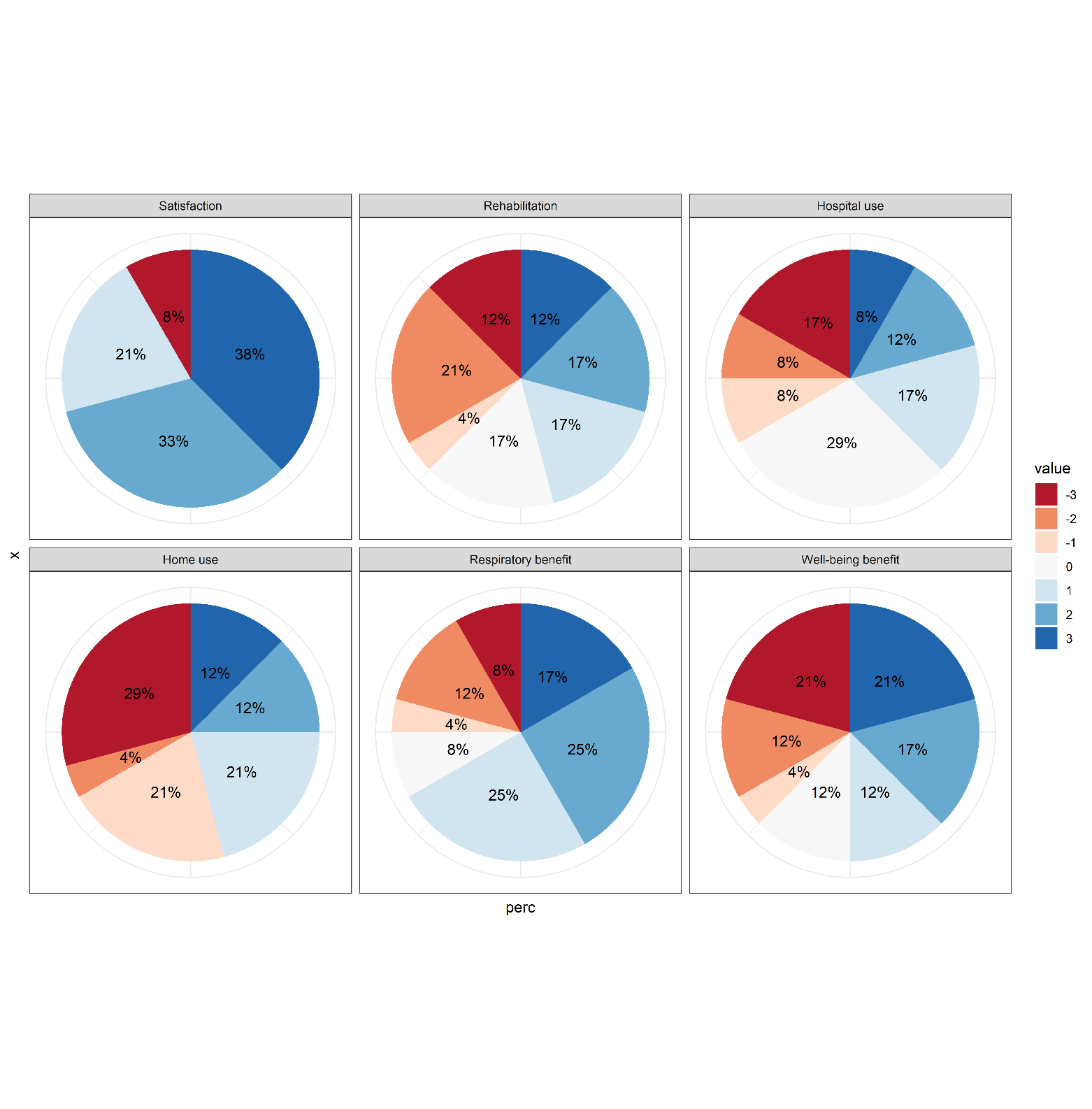

Figure S2 Percentage of feasibility scores for all items, rated on a 7-point Likert scale, with -3 = Strongly disagree, -2 = Disagree; -1 = Somewhat disagree; 0 = Neither agree nor disagree; 1 = Somewhat agree; 2 = Agree; 3= Strongly agree
